# Toward precision rehabilitation in adolescent mild traumatic brain injury: leveraging physiologic data from commercially available smartwatches to identify patient subgroups

**DOI:** 10.64898/2026.07.16.26358245

**Authors:** Sarah A. Kettlety, Emily R. Akrong, Stacy J. Suskauer, Ryan T. Roemmich, Beth S. Slomine, Adrian M. Svingos

**Affiliations:** Brain Injury Clinical Research Center, Kennedy Krieger Institute, Baltimore, MD, USA; Department of Physical Medicine and Rehabilitation, Johns Hopkins University School of Medicine, Baltimore, MD, USA; Center for Movement Studies, Kennedy Krieger Institute, Baltimore, MD, USA; Department of Psychiatry and Behavioral Sciences, Johns Hopkins University School of Medicine, Baltimore, MD, USA

**Author notes:** These authors contributed equally to this work (Sarah A. Kettlety, Emily R. Akrong).

**Keywords:** mild traumatic brain injury, concussion, adolescents, autonomic dysfunction, Fitbit, remote monitoring, clustering, physical activity

## Abstract

**Objective:** To identify meaningful subgroups of adolescents with mTBI using physiologic and physical activity data obtained via consumer-grade smartwatches.

**Setting:** Specialty concussion clinic.

**Participants:** Eighty participants aged 13-18 within six months of mTBI diagnosis were enrolled. Sixty-one participants were included in the analysis.

**Design:** Prospective longitudinal cohort study. Participants wore a Fitbit Sense 2. Heart rate and step count data collected within fourteen days of enrollment were included.

**Main measures:** A group-based steps per minute (SPM) threshold (25th percentile; 10 SPM) and individualized heart rate threshold (20% heart rate reserve (HRR)) were used to classify each minute of active daytime data into quadrants: SPM>10 & HRR>20% (QI), SPM≤10 & HRR>20% (QII), SPM≤10 & HRR≤20% (QIII), and SPM>10 & HRR≤20% (QIV). Percentage of minutes in QI, QII, and QIV, mean steps per day, percentage of minutes with zero steps, mean SPM in QI, and resting heart rate were included in a k-means clustering algorithm. Subgroup differences by clustering variables were evaluated using Kruskal-Wallis tests.

**Results:** Three subgroups emerged: Sedentary (n=12), Active (n=23), and Atypically Elevated Heart Rate (AEHR; n=26). Subgroups varied significantly on all clustering variables (*P*<0.01). The Active subgroup took a high number of steps per day, had lower sedentary time, and had the highest mean SPM in QI. The Sedentary subgroup took fewer steps per day than the Active subgroup, had high sedentary time, and showed the highest resting heart rate. The AEHR subgroup took fewer steps per day than the Active subgroup, had high sedentary time, and spent a higher percentage of time with an atypically high heart rate response to low levels of activity than the other subgroups.

**Conclusions:** Data from wearable devices can identify subgroups of adolescents with mTBI with distinct physiologic/physical activity profiles, which may ultimately be used to inform personalized activity prescriptions.

## INTRODUCTION

Autonomic dysfunction is a common sequela of mild traumatic brain injury (mTBI),^1,2^ including orthostatic tachycardia and orthostatic hypotension.^3–5^ Autonomic dysfunction is associated with exercise intolerance and symptoms that interfere with functioning including lightheadedness, dizziness, headache, blurry vision, and brain fog.^1,6^ Previous studies have reported that ~10% of youth with mTBI demonstrate objective indicators of autonomic dysfunction.^3,4^

Physical activity prescription is an important method for clinical management of autonomic dysfunction^6,7^ and mTBI symptoms.^8,9^ Early sub-symptom threshold physical activity following mTBI results in faster recovery and lowers risk for persisting symptoms.^9–11^ However, symptoms of autonomic dysfunction (e.g., lightheadedness, tachycardia) may cause a patient to avoid exercise. Reduced physical activity, particularly during the initial weeks post-injury, can further exacerbate autonomic dysfunction and delay recovery.^8^ Early identification of autonomic dysfunction may lead to tailored physical activity prescription (e.g., recumbent activities, initially short daily bouts of aerobic activity) and/or incorporation of other interventions to manage symptoms (e.g., compression, hydration, medications).^6^

Management of mTBI involves assessing patients’ level of physical activity and degree of associated symptom exacerbation to guide activity progression. However, self-and caregiver-reported level of activity may not accurately reflect — or adequately characterize — patients’ level of physical activity.^12^ Consumer-grade wearable devices, such as the Fitbit device^13,14^ represent an opportunity for objective tracking of physical activity data, including via minute-level step count and heart rate data. In addition to understanding physical activity level, clinicians also benefit from obtaining data on physiologic response activity (i.e., heart rate).

Point-in-time exertional assessments, such as the Buffalo Concussion Treadmill/Bike Tests,^15,16^ provide valuable insight on physiologic response to high intensity activity and inform exercise prescription. However, point-in-time exertional assessments require time and resources that may not be feasible in all clinical settings. Additionally, adolescents with mTBI may experience orthostatic tachycardia,^3,4^ which can appear as atypically high heart rate during relatively low levels of activity that would not be captured by exertional assessments alone. Using a Fitbit device provides an opportunity to collect physiologic and physical activity data captured across a range of activity intensities and provide insight into real-world functioning.

Previous work has used heart rate and step count data from Fitbit devices to subgroup individuals post-stroke.^17^ Authors identified three distinct subgroups of individuals post-stroke: Active, Deconditioned, and Sedentary. Importantly, both the Deconditioned and Sedentary subgroups took relatively few steps per day, but the heart rate response to activity differed.^17^ The Deconditioned group spent a significantly greater percentage of time with an elevated heart rate response to low levels of activity.^17^ This subgrouping method may be particularly informative in the context of adolescent mTBI, as we expect a subgroup of patients to experience autonomic dysfunction.

Here, we leveraged commercially available wearable technology (i.e., Fitbits) to obtain real-world, objective data on the interplay between physical activity (steps) and physiologic response to activity (heart rate). We aimed to identify subgroups of adolescents with mTBI with distinct physiologic responses to activity using heart rate and step count data collected from a Fitbit device. We hypothesized that we would identify distinct subgroups using heart rate and step count data, with one subgroup demonstrating an atypical heart rate response to minimal activity.

To better understand whether the subgroups align with clinically available data, we also explored the relationship between the subgroups and demographic and clinical data.

## METHODS

### Participants

Adolescents diagnosed with mTBI^18^ were recruited from the Kennedy Krieger Multidisciplinary Concussion Clinic from March 2024 to June 2025. Electronic medical charts were reviewed for patients aged 13-18 years and within 6 months of injury. Patients were not approached for screening by the study team if they met any of the following exclusion criteria: pre-existing functional limitations (e.g., intellectual disability), foster care status, or the presenting parent/caregiver was not fluent in English. The patient, caregiver, or clinician could also opt the patient out of screening. A study team member screened identified patients in person during their scheduled appointment. Patients were included if they were willing to wear an activity monitor for a year, had in-home Wi-Fi, and had a personal smart phone. The Johns Hopkins Medicine Institutional Review Board approved this protocol (IRB00247292). We obtained oral assent/parental consent from participants aged 13-17 years. Oral consent was obtained from participants aged 18. A priori power analysis required n=20 per expected subgroup.^19^ We enrolled 80 participants to account for possible attrition over the study period.

### Experimental procedures

Participant sex, race, ethnicity, history of previous mTBI, and pre-injury sports participation^20^ were reported by participants and their caregivers. The following items were abstracted from the medical charts: mTBI injury mechanism, days from mTBI to enrollment, and history of neurodevelopmental, mental health, and/or autonomic (e.g. postural orthostatic tachycardia syndrome, orthostatic intolerance) diagnoses.

To capture symptom burden, participants completed the Generalized Anxiety Disorder-7 (GAD-7),^21^ Patient Health Questionnaire-8 (PHQ-8),^22^ and Rivermead Post-Concussion Symptoms Questionnaire (RPQ)^23^ at enrollment. Participants completed the GAD-7 and PHQ-8 twice: once reporting symptoms two weeks prior to their injury (pre-injury symptoms) and once reporting symptoms since the injury (post-injury symptoms). The RPQ was completed once, as it asks the participant to compare the symptoms they experienced in the past 24 hours to what they experienced before the injury. Detailed descriptions of the questionnaires and scoring can be found in the Supplemental Methods (see Supplemental Digital Content 1). All demographic and clinical data were collected and managed using the REDCap electronic data capture tools.^24,25^

The Fitbit data collection procedures have been described previously.^13^ All participants were given a Fitbit Sense 2 device. Participants were asked to wear the device for one year on their preferred wrist. For this study, we considered minute-level heart rate and step count data collected during the first fourteen days following device setup.

### Data analysis

To ensure wear time adequate to estimate daily physical activity, we first identified valid days. We considered a day valid if at least one step was recorded and minute-level heart rate data were recorded for at least 75% of daytime hours (7:00AM-9:59PM).^13,17^ For each participant, we sorted the valid days by the greatest number of valid minutes, then by chronological day order.

We then selected the first seven days with the highest number of valid minutes. Participants were required to have seven valid days to be included in the analyses (these seven days could be non-consecutive).

We then categorized each minute of daytime activity for each participant into one of four quadrants using a step count and heart rate threshold (QI-QIV; Table 1). The step count threshold was defined as the 25^th^ percentile of the group steps per minute distribution (of minutes with at least one step recorded).^17^ The heart rate threshold was defined for each participant as 20% heart rate reserve (HRR; equation 1).^26^ We used the Tanaka equation (equation 2)^27^ to calculate each participant’s maximum heart rate (HR_max_).^28^ We calculated resting heart rate (HR_rest_) during nighttime hours (12:00pm-6:59am) across two valid nights. Specific details about the HR_rest_ calculation can be found in the Supplemental Methods (see Supplemental Digital Content 1).^29^

**Table 1.** Quadrants used to describe heart rate response to activity.

| Quadrant | Heart rate | Steps | Interpretation |
| --- | --- | --- | --- |
| QI | > 20 % HRR | > 25<sup>th</sup> percentile | Expected heart rate response to activity |
| QII | > 20 % HRR | ≤ 25<sup>th</sup> percentile | Atypically elevated heart rate response to minimal activity |
| QIII | ≤ 20 % HRR | ≤ 25<sup>th</sup> percentile | Expected heart rate response to minimal activity |
| QIV | ≤ 20 % HRR | > 25<sup>th</sup> percentile | Conditioned heart rate response to activity |

The heart rate threshold was individualized. The group-based steps per minute threshold was defined as the 25^th^ percentile of group steps per minute distribution. Abbreviations: QI, quadrant I; QII, quadrant II; QIII, quadrant III; QIV, quadrant IV; HRR, heart rate reserve.

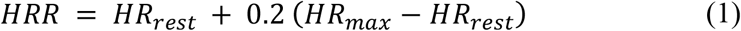

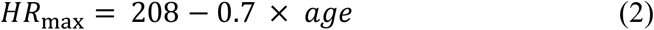

### Statistical analysis

All analyses were performed in R version 4.5.2.^30^ We used k-means clustering to identify subgroups of mTBI patients using the kmeans function.^17^ We used seven variables in the k-means analysis: Percentages of time spent in QI, QII, and QIV, percentage of daytime minutes with 0 steps (i.e., sedentary time), average steps per day, average number of steps per minute in QI, and resting heart rate.^17^ All variables were scaled (mean = 0, standard deviation = 1) before clustering. To select the number of clusters, we used the NbClust package which filters through thirty indices to determine the optimal number of clusters.^31^ We assessed cluster stability using the Jaccard coefficient, calculated using the clusterboot function.^32^ Clusters with an average Jaccard coefficient value > 0.75 were considered stable.^33^

We next examined the differences in all clustering variables between subgroups. We utilized Kruskal-Wallis tests, using Dunn’s test as a post-hoc test on statistically significant variables.

To understand the relative importance of each clustering variable in cluster assignment, we used a random forest algorithm. We used subgroup assignment as the outcome variable and used the importance function in the randomForest package to assess the order of variable importance.^34^

Finally, we explored the differences in demographic and mTBI characteristics by subgroup. For continuous variables, we utilized Kruskal-Wallis tests with Dunn’s post-hoc tests. For categorical variables, we used Fisher’s Exact tests with pairwise Fisher’s Exact post-hoc tests. We compared subgroups by age, sex, race, ethnicity, and medical history. We also compared subgroups by injury mechanism, days since injury, presence of post-traumatic amnesia, loss of consciousness, pre-injury sports participation, pre-injury GAD-7, pre-injury PHQ-8, post-injury GAD-7, post-injury PHQ-8, and RPQ total score. Of note, one participant was missing data from the post-injury GAD-7 and was removed from the post-injury GAD-7 analysis. Four participants missed one item on the RPQ, and one participant missed two items. We addressed this by imputing each participant’s average score across all answered items and included that value as the missing item(s) score in the RPQ total score.^35^

## RESULTS

Eighty participants were enrolled (Figure 1). One participant never synced their Fitbit device, one participant withdrew prior to initiating data collection, and three participants’ Google accounts were suspended. Fourteen participants were excluded from the analyses due to not having seven valid days and two valid nights of data. Thus, 61 participants were included in the analyses.

**Figure 1.**
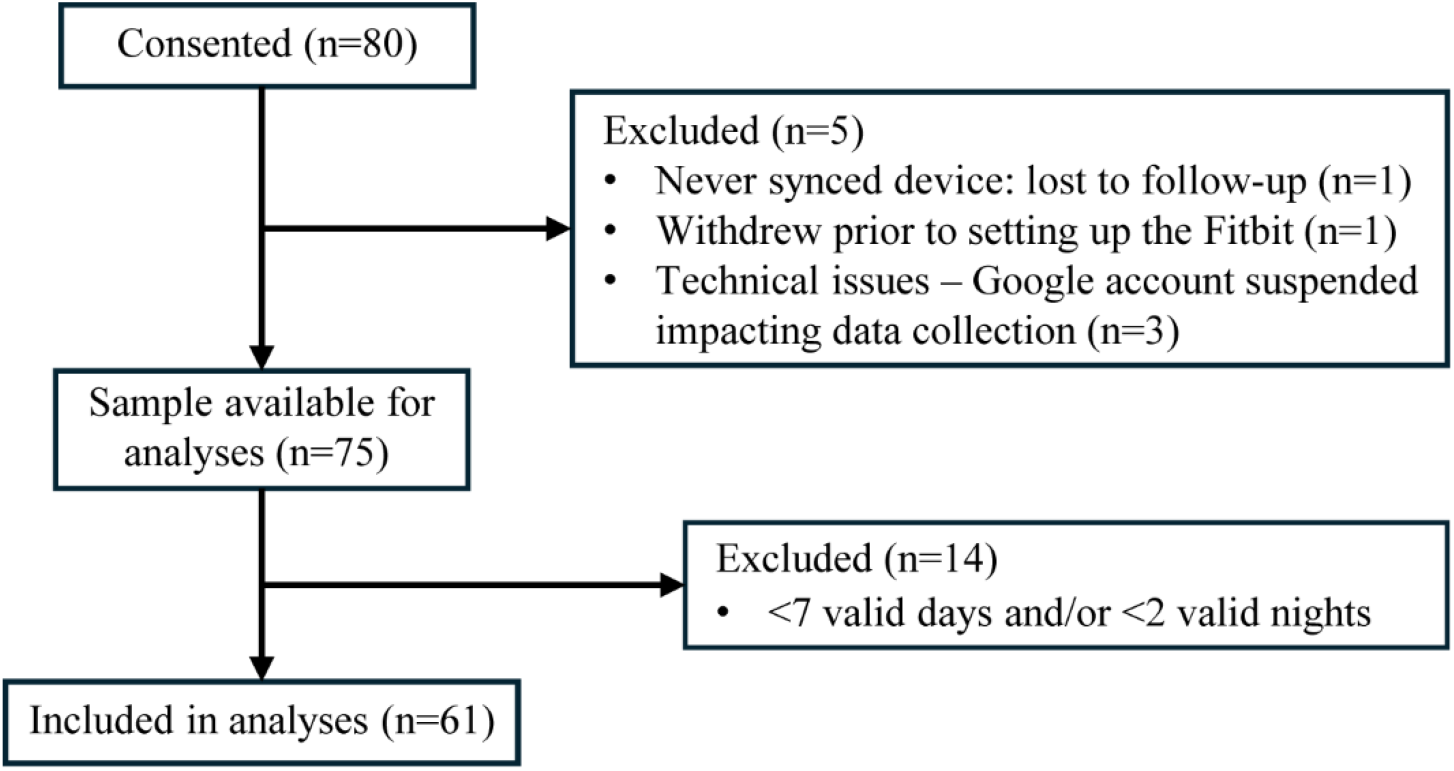
Flow Diagram of participant enrollment and sampling for analyses.

### Step count and heart rate thresholds

The 25th percentile of the group-based steps per minute distribution was 10 steps per minute and acted as our step count threshold. The individualized heartrate thresholds (20% HRR) ranged from 73-96 bpm.

### Subgroups based on clustering variables

Three distinct subgroups emerged. We named the subgroups “Sedentary” (n=12), “Atypically Elevated Heart Rate (AEHR)” (n=26), and “Active” (n=23) based on their clustering variable profiles (Figure 2). Mean Jaccard Coefficients for the subgroups were 0.75 (Sedentary), 0.81 (AEHR), and 0.77 (Active), indicating stability for all clusters.

**Figure 2.**
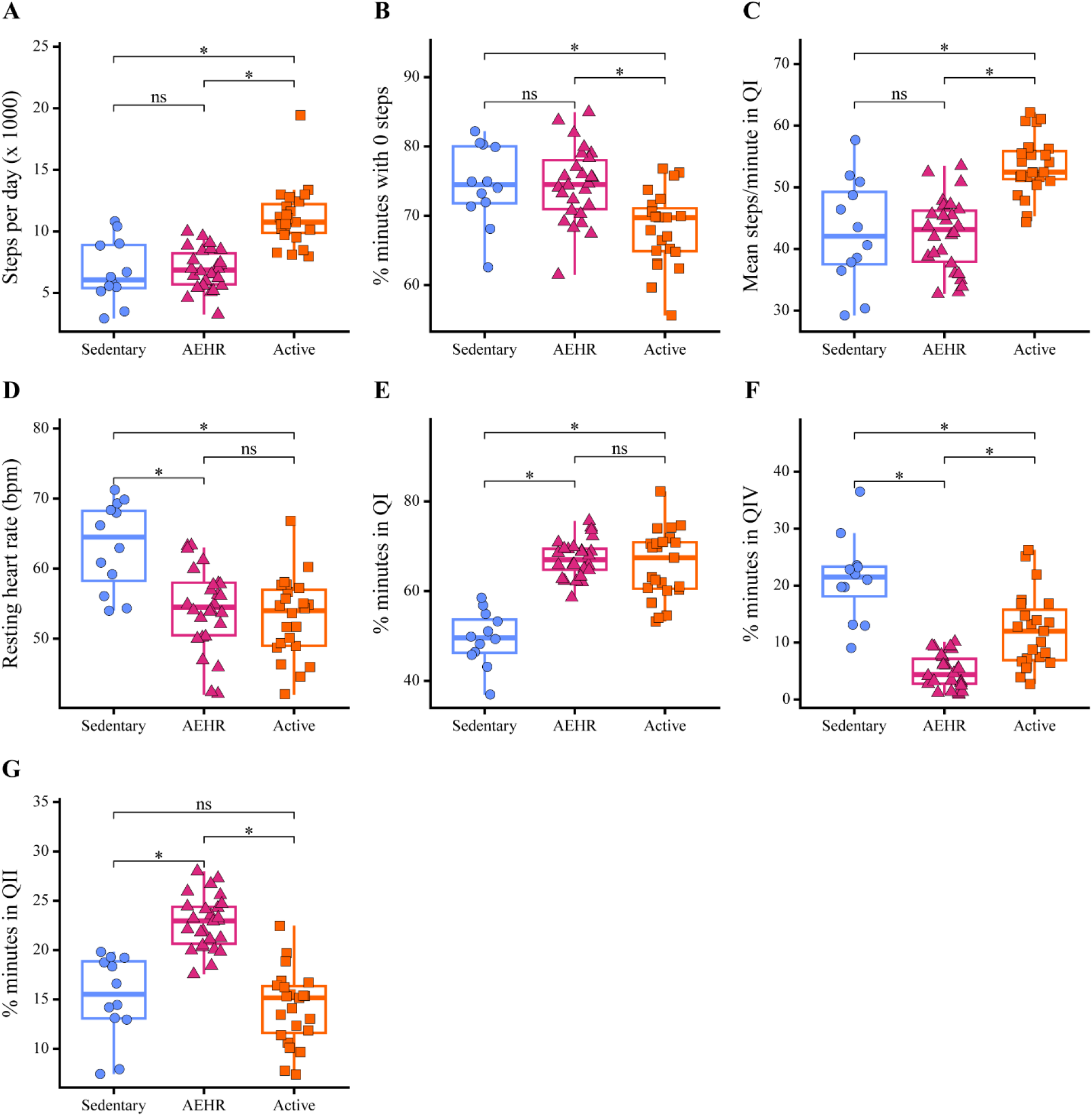
Comparison of subgroups by steps per day (A), percentage of minutes with zero steps (B), mean steps per minute in quadrant I (C), resting heart rate (D), percentage of minutes in quadrant II (E), percentage of minutes in quadrant IV (F), and percentage of minutes in quadrant II (G). The boxplots show subgroup medians with upper and lower quartiles. Individual participant data is represented by the symbols. *Significant pairwise difference at p<0.05. Abbreviations: AEHR, atypically elevated heart rate; ns, not significant; QI, quadrant I; QII, quadrant II; QIV, quadrant IV.

We found significant differences among subgroups for all clustering variables (see Supplemental Digital Content 1, Supplemental Table 1 for subgroup medians and model results). Specific differences between subgroups identified with post-hoc testing are described below and displayed in Figure 2.

The Active subgroup took the highest number of steps per day compared to the Sedentary and AEHR subgroups (both *P*<0.001, Figure 2A). The Active subgroup spent a smaller percentage of minutes with zero steps than the AEHR and Sedentary subgroups (*P*=0.001 and *P*=0.008, respectively; Figure 2B). The Active subgroup also had the highest steps per minute in QI, indicating higher intensity activity in QI compared to the other subgroups (both *P*<0.001; Figure 2C).

The Sedentary subgroup had a higher resting heart rate compared to the Active and AEHR subgroups (both *P*<0.001; Figure 2D). The Sedentary subgroup also spent the lowest percentage of minutes in QI compared to the other subgroups (both *P*≤0.001; Figure 2E), reflecting less time engaging in light to heavy activity throughout the day. The Sedentary subgroup had the highest percentage of minutes in QIV compared to the Active and AEHR subgroups (*P*=0.02 and *P*<0.001, respectively; Figure 2F).

The AEHR subgroup had high sedentary time (percentage of minutes with zero steps) like the Sedentary subgroup (*P*=0.97; Figure 2B). The AEHR subgroup also spent the greatest percentage of minutes in QII compared to the Sedentary and Active subgroups (both *P*<0.001, Figure 2G). This increased time in QII reflects more time spent with an atypical heart rate response to a minimal amount of activity. The AEHR subgroup overall spent the same percentage of minutes in QI as the Active subgroup (*P*=0.62; Figure 2E). However, the AEHR subgroup took fewer steps per minute in QI (*P*<0.001; Figure 2C), indicating reduced intensity.

The AEHR subgroup also spent the lowest percentage of minutes in QIV compared to the Active and Sedentary subgroups (both *P*<0.001; Figure 2F). These results suggest that participants in the AEHR subgroup have a high heart rate response to minimal activity, with most of their active minutes spent above the individual heart rate threshold.

Random forest analysis revealed percent of minutes in QII as the most critical variable in determining subgroup assignment, followed by percent minutes in QI (Figure 3).

**Figure 3.**
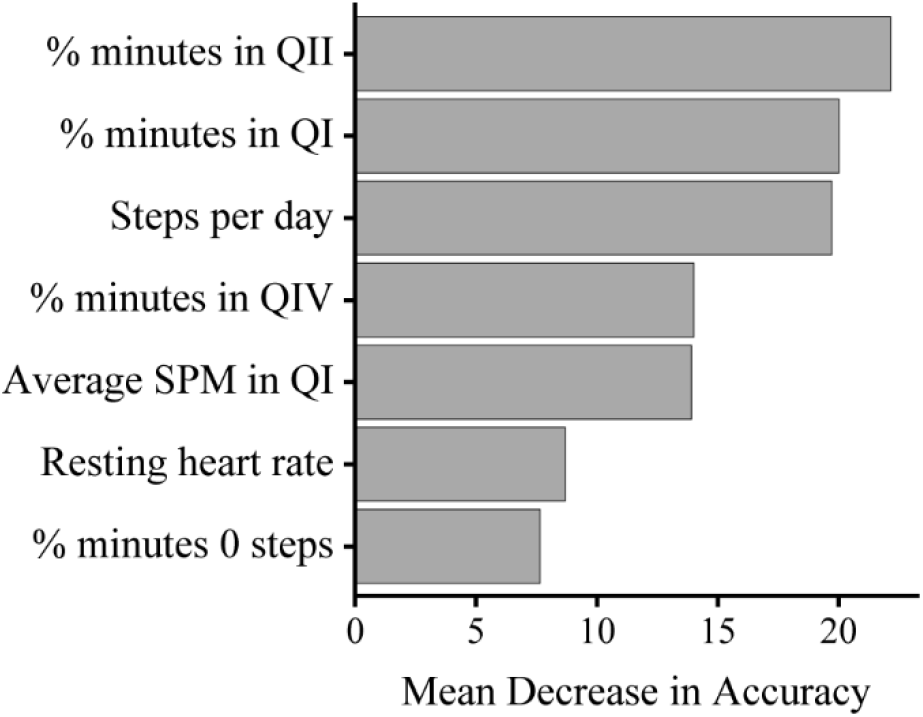
Variable importance results from the random forest analysis. Abbreviations: QII, quadrant II; QI, quadrant I; QIV, quadrant IV, SPM, steps per minute.

### Demographic and mTBI characteristics by subgroup

Table 2 summarizes participant demographic, medical history, and mTBI characteristics by subgroup. The subgroups did not differ on age at enrollment (H(2)=1.2, *P*=0.55), race (*P*=0.11), or ethnicity (*P*=0.90). We found a significant difference in sex across subgroups (*P*<0.001), with more males in the Active subgroup than in the Sedentary (*P*<0.001) and the AEHR (*P*=0.006) subgroups. There was no significant difference in sex between the Sedentary and AEHR subgroups (*P*=0.29). The subgroups also differed in history of a prior mTBI requiring more than one week to recover (*P*=0.03). More participants in the AEHR subgroup had a prior mTBI requiring more than one week to recover compared to the Sedentary subgroup (*P*=0.03). The percentage of participants with a prior mTBI with recovery more than one week did not differ between the AEHR and Active subgroups (*P*=0.14) or Active and Sedentary subgroups (*P*=0.14).

**Table 2.** Participant demographics, medical history, and mTBI characteristics.

|  | **Overall**<br>**(n=61)** | **Sedentary**<br>**(n=12)** | **AEHR**<br>**(n=26)** | **Active**<br>**(n=23)** | **P-value** |
| --- | --- | --- | --- | --- | --- |
| Age (years), median (IQR) | 15.7 (14.5-16.8) | 15.3 (14.5-15.9) | 16.0 (14.5-16.9) | 15.5 (14.3-16.7) | 0.55 |
| Sex, n (%) |  |  |  |  | <0.001* |
| Female | 44 (72.1%) | 12 (100%) | 22 (84.6%) | 10 (43.5%) |  |
| Male | 17 (27.9%) | 0 (0.0%) | 4 (15.4%) | 13 (56.5%) |  |
| Race, n (%) |  |  |  |  | 0.11 |
| White | 40 (65.6%) | 5 (41.7%) | 19 (73.1%) | 16 (69.6%) |  |
| Black | 8 (13.1%) | 4 (33.3%) | 1 (3.8%) | 3 (13.0%) |  |
| Asian | 1 (1.6%) | 0 (0.0%) | 1 (3.8%) | 0 (0.0%) |  |
| More than one | 9 (14.8%) | 3 (25.0%) | 4 (15.4%) | 2 (8.7%) |  |
| Other | 3 (4.9%) | 0 (0.0%) | 1 (3.8%) | 2 (8.7%) |  |
| Ethnicity, Hispanic/Latino, n (%) | 9 (14.8%) | 1 (8.3%) | 4 (15.4%) | 4 (17.4%) | 0.90 |
| Prior mTBI, n (%) | 27 (44.3%) | 3 (25.0%) | 14 (53.8%) | 10 (43.5%) | 0.28 |
| Prior mTBI >1 week recovery, n (%) | 20 (32.8%) | 1 (8.3%) | 13 (50.0%) | 6 (26.1%) | 0.03* |
| ADHD, n (%) | 15 (24.6%) | 3 (25.0%) | 6 (23.1%) | 6 (26.1%) | >0.99 |
| SLD, n (%) | 7 (11.5%) | 3 (25.0%) | 2 (7.7%) | 2 (8.7%) | 0.30 |
| Autonomic, n (%) | 4 (6.6%) | 2 (16.7%) | 1 (3.8%) | 1 (4.3%) | 0.32 |
| Mental Health dx, n (%) | 29 (47.5%) | 7 (58.3%) | 10 (38.5%) | 12 (52.2%) | 0.48 |
| Injury Mechanism, n (%) |  |  |  |  | 0.18 |
| Sports-related | 38 (62.3%) | 9 (75.0%) | 15 (57.7%) | 14 (60.9%) |  |
| Automobile-related | 11 (18.0%) | 0 (0.0%) | 6 (23.1%) | 5 (21.7%) |  |
| Non-auto fall/accident | 4 (6.6%) | 1 (8.3%) | 1 (3.8%) | 2 (8.7%) |  |
| Assault | 3 (4.9%) | 1 (8.3%) | 0 (0.0%) | 2 (8.7%) |  |
| Other | 5 (8.2%) | 1 (8.3%) | 4 (15.4%) | 0 (0.0%) |  |
| Days since injury, median (IQR) | 18 (11-35) | 24 (14-51) | 19 (10-31) | 18 (20-35) | 0.58 |
| Post-traumatic amnesia, n (%) | 19 (31.1%) | 2 (16.7%) | 10 (38.5%) | 7 (30.4%) | 0.72 |
| Loss of consciousness, n (%) | 13 (21.3%) | 1 (8.3%) | 8 (30.8%) | 4 (17.4%) | 0.12 |
| RPQ total, median (IQR) | 19 (10-27) | 19 (17-24) | 20 (14-30) | 16 (5-23) | 0.29 |
| Pre-injury GAD-7, median (IQR) | 3 (0-5) | 4 (1-7) | 3 (1-6) | 2 (0-5) | 0.45 |
| Post-injury GAD-7, median (IQR) | 4 (2-7) | 5 (4-7) | 5 (2-8) | 3 (0-5) | 0.05 |
| Pre-injury PHQ-8, median (IQR) | 2 (1-5) | 3 (1-6) | 2 (0-5) | 2 (0-4) | 0.47 |
| Post-injury PHQ-8, median (IQR) | 6 (4-10) | 8 (5-12) | 6 (4-10) | 6 (3-9) | 0.31 |
| Pre-injury sports participation, n (%) |  |  |  |  | >0.99 |
| School or travel/select | 47 (77.0%) | 9 (75.0%) | 20 (76.9%) | 18 (78.3%) |  |
| Recreational or none | 14 (23.0%) | 3 (25.0%) | 6 (23.1%) | 5 (21.7%) |  |

The subgroups also did not differ on days since injury (H(2)=1.1, *P*=0.58), presence of post-traumatic amnesia (*P* = 0.72), or loss of consciousness (*P*=0.12). We failed to detect a significant difference between groups by pre-injury GAD-7 (H(2)=1.6, *P*=0.45), pre-injury PHQ-8 (H(2)=1.5, *P*=0.47), post-injury GAD-7 (H(2)=5.9, *P*=0.05), or post-injury PHQ-8 (H(2)=2.4, *P*=0.31). Total RPQ scores also did not differ by subgroup (H(2)=2.5, *P*=0.29).

We utilized Kruskal-Wallis tests for continuous variables and Fisher’s Exact tests for categorical variables. Abbreviations: AEHR, atypically elevated heart rate; mTBI, mild traumatic brain injury; ADHD, Attention-Deficit/Hyperactivity Disorder; SLD, Specific Learning Disability; RPQ, Rivermead Post-Concussion Symptoms Questionnaire; GAD-7, General Anxiety Disorder-7; PHQ-8, Patient Health Questionnaire-8.

## DISCUSSION

In this study, we aimed to identify subgroups of adolescents presenting for specialty care after diagnosed mTBI using heart rate and step count data collected from a Fitbit device. We identified three subgroups: Active, Sedentary, and AEHR. Each subgroup displayed a distinct physiologic/physical activity profile that may provide a foundation for personalizing rehabilitation in adolescents with mTBI. The Active subgroup took a high number of steps per day, had lower sedentary time, and had the highest activity intensity. The Sedentary subgroup took fewer steps per day, had high sedentary time, and had the highest resting heart rate. The AEHR subgroup took fewer steps per day and had high sedentary time (similar to the Sedentary subgroup). The AEHR subgroup also spent a greater percentage of time with an atypically high heart rate response to low levels of activity than the other subgroups, and this was the most important variable in determining subgroup classification.

This work highlights the potential for using real-world physiologic data to subgroup adolescents after mTBI, which could ultimately lead to more precisely targeted rehabilitation approaches. For example, the AEHR subgroup may reflect a subset of patients with autonomic dysfunction that may need guidance on how to exercise within their symptom tolerance (e.g., using a recumbent bike) or benefit from a referral to physical therapy.^6^ In contrast, adolescents in the Sedentary subgroup, who are inactive but largely demonstrate an expected response to activity, may benefit from a behavioral intervention focused on how to initiate and maintain an exercise routine.

Focusing only on increasing activity in those experiencing atypical physiologic responses may cause symptom exacerbation when trying to exercise, which may lead to reduced activity and delayed recovery.^8^

Our work extends prior research^17^ using step count and heart rate data to identify subgroups in clinical populations. Koffman et al.^17^ identified three subgroups of adults with stroke: Active, Sedentary, and Deconditioned. The participants post-stroke in the Deconditioned subgroup spent significantly more time with an exaggerated heart rate response to minimal activity compared to the other subgroups,^17^ similar to the AEHR subgroup found in our work. Both the Deconditioned subgroup of adults with stroke and the AEHR subgroup of adolescents with mTBI demonstrated a physiologically disproportionate heart rate response to minimal activity. Together, these studies highlight the importance of considering both step count and heart rate in conjunction, as examining step count alone in either of these populations would have missed the heightened physiologic responses to activity, which may be of clinical importance.

It is possible that the adolescents with mTBI may be deconditioned (as is hypothesized in adults post-stroke^17^). However, there are a few reasons this is less likely in our sample of adolescents post-mTBI. Resting heart rate in the AEHR subgroup did not differ from the Active subgroup. If the physiologic changes in the AEHR subgroup were primarily due to deconditioning, we would also expect the AEHR subgroup to have a higher resting heart rate. Other factors that could contribute to deconditioning, such as time since injury and pre-injury sports participation,^36^ were not different between subgroups, contributing to our understanding that this may represent post-mTBI exercise intolerance as opposed to deconditioning.^37^

We found that the percentage of females across subgroups differed. The Active subgroup had a similar percentage of males and females, whereas the Sedentary and AEHR subgroups were predominately female. Females are more likely to present with autonomic dysfunction related diagnoses, such as postural orthostatic tachycardia syndrome,^38^ which aligns with our findings.

However, orthostatic intolerance resulting from mTBI typically does not show a prevalence difference between sexes.^3–5,39,40^ The greater percentage of females in the AEHR subgroup may be due in part to the female predominance of our sample. We are also inferring the presence of autonomic dysfunction using Fitbit data, and did not use head-upright tilt table or standing tests to characterize orthostatic intolerance.^3–5,39,40^

We also found that the percentage of participants with a prior mTBI requiring more than one week to recover differed between subgroups. The percentage of participants in the AEHR subgroup was greater than the Sedentary subgroup. This aligns with previous work that found that 79% of participants with orthostatic intolerance had at least one previous mTBI.^4^ A prior mTBI requiring more than one week to recover also increases the likelihood of persisting symptoms after mTBI.^41^ The percentage of participants with a prior mTBI requiring more than one week to recover did not differ between the AEHR and Active subgroups. This may suggest that participants with a prior mTBI requiring more than one week to recover who are more physically active may recover autonomic function more quickly or may be less likely to experience autonomic dysfunction after mTBI. However, we are unable to directly test this hypothesis in this work.

We did not find differences in anxiety, depression, or overall symptom burden between subgroups. The lack of alignment with typical symptom scales suggests that the subgrouping may be describing unique information related to the physiologic/physical activity profile that is not captured by traditional measures. Though we did not find a statistical difference in anxiety between subgroups, future research should continue to examine the role of anxiety as it commonly co-occurs with autonomic dysfunction,^42^ and is related to exercise intolerance in adults after mTBI.^43^ The symptom measures used in this work were also retrospective self-reports, which may differ from symptoms over the two-week period the Fitbit data was collected or symptoms under physical and/or mental exertion. Our results align with previous findings showing that symptoms did not differ between those who exhibited objective indicators of autonomic dysfunction and those who did not.^3,4,44^ Similarly, other work found that screening for common orthostatic intolerance symptoms did not help identify patients with orthostatic intolerance, with only 29% of symptomatic patients having positive standing test findings.^5^

### Limitations and future directions

Our study had several limitations which can be addressed through future research. First, we were powered to find three clusters.^19^ Though our clusters were stable, it is possible that with greater sample sizes we may identify more subgroups. Future work should investigate if different subgroups emerge with larger sample sizes. Second, we did not collect data from standing tests, head-upright tilt table tests, COMPASS-31 survey,^45^ or point-in-time exertional assessments.

While we found indications of atypical physiologic responses to activity in our subgroups using real-world Fitbit data, we do not currently understand how the subgroups relate to other measures of autonomic dysfunction and exercise intolerance. Future work should investigate the relationship between subgroups and these measures. Finally, we performed a cross-sectional clustering analysis with data collected within fourteen days of enrollment. Future work with larger sample sizes should investigate how subgroups change beyond clinical recovery using longitudinal clustering approaches, and how the subgroups may relate to long-term outcomes such as persisting symptoms after mTBI or onset of new diagnoses.

### Conclusion

We leveraged physiologic and physical activity data from commercially available wearable devices to identify three distinct subgroups of adolescents with mTBI, including one subgroup characterized by an atypically elevated heart rate response to minimal activity. This method may ultimately be used to inform personalized rehabilitation approaches in adolescents with mTBI. Future work should aim to understand how subgroups relate to recovery trajectories and if they may help identify those at risk for adverse health outcomes following mTBI.

## Supporting information

Supplemental Digital Content 1

## Data Availability

The datasets from this study are available from the senior author (AMS) on reasonable request.

## Acknowledgments

The authors acknowledge Lily Koffman for sharing the analysis code used in her previous work, the Johns Hopkins Technology Innovation Center for the development of the application used to collect the Fitbit data, and the Kennedy Krieger Institute and Johns Hopkins University School of Medicine Precise Center Team for their support of this work.

