## Supplemental Digital Content 1 for "Toward precision rehabilitation in adolescent mild traumatic brain injury: leveraging physiologic data from commercially available smartwatches to identify patient subgroups"

### SUPPLEMENTAL METHODS

*Symptom questionnaires*

The GAD-7 provides seven statements and asks the participant how often they experienced the feelings described in each statement over the past two weeks, ranging from 0 (not at all) to 3 (nearly every day), for a maximum total score of 21. The PHQ-8 lists eight problems and asks the participant to rate how often they have been bothered by each problem over the past two weeks. For each item, scores range from 0 (not at all) to 3 (nearly every day), for a maximum score of 24. The RPQ lists sixteen common post-concussive symptoms and asks the participant to compare the symptoms they experienced in the past 24 hours to what they experienced before the injury. The ratings for each symptom range from 0 (not experienced) to 4 (severe problem). We scored items with a rating of 1 (no more of a problem) as 0, for a total possible score out of 64.[1]

*Resting heart rate calculation.*

We calculated resting heart rate ($HR_{rest}$) during nighttime hours (12:00pm-6:59am). To do this, we first identified minutes with a recorded heart rate and with step count equal to zero (i.e., valid minutes). We did this to ensure we were using heart rate data during periods of rest and not activity, as sleep disturbances are common post-mTBI.[2] We then identified valid nights as nights with at least 315 valid minutes (75%). For each participant, we sorted the valid nights by the greatest number of valid minutes, then by chronological night order. We then selected the first two nights with the highest number of valid minutes. Therefore, participants were required to have two valid nights (in addition to seven valid days) to be included. Finally, we calculated a five-minute rolling average across the two nights for each participant.[3] Resting heart rate was defined as the first percentile value from the five-minute rolling average.[3]

### SUPPLEMENTAL RESULTS

**Supplemental Table 1.** Kruskal-Wallis and Dunn's post-hoc results for clustering variables.

| | **Active** Median (IQR) | **AEHR** Median (IQR) | **Sedentary** Median (IQR) | **Kruskal-Wallis H** | **Kruskal-Wallis p-value** | **Dunn's post-hoc p-values** |
|---|---|---|---|---|---|---|
| Steps per day | 10745 (9883-12218) | 6858 (5705-8223) | 6064 (5402-8897) | 31.7 | <0.001* | Active vs. AEHR: <0.001<br>Active vs. Sedentary: <0.001<br>Sedentary vs. AEHR: 0.91 |
| % of minutes with 0 steps | 70% (65%-71%) | 75% (71%-78%) | 74% (71%-80%) | 15.0 | <0.001* | Active vs. AEHR: 0.001<br>Active vs. Sedentary: 0.008<br>Sedentary vs. AEHR: 0.97 |
| Average steps per minute in QI | 53 (51-55) | 43 (38-46) | 42 (38-49) | 27.7 | <0.001* | Active vs. AEHR: <0.001<br>Active vs. Sedentary: <0.001<br>Sedentary vs. AEHR: 0.73 |
| Resting heart rate (bpm) | 54 (49-57) | 55 (51-58) | 65 (58-68) | 15.3 | <0.001* | Active vs. AEHR: 0.50<br>Active vs. Sedentary: 0.001<br>Sedentary vs. AEHR: <0.001 |
| % minutes in QI | 67% (61%-71%) | 67% (65%-69%) | 50% (46%-54%) | 26.6 | <0.001* | Active vs. AEHR: 0.62<br>Active vs. Sedentary: <0.001<br>Sedentary vs. AEHR: <0.001 |
| % minutes in QII | 15% (12%-16%) | 23% (21-24%) | 16% (13%-19%) | 39.6 | <0.001* | Active vs. AEHR: <0.001<br>Active vs. Sedentary: 0.60<br>Sedentary vs. AEHR: <0.001 |
| % minutes in QIV | 12% (7%-16%) | 4% (3%-7%) | 22% (18%-23%) | 33.9 | <0.001* | Active vs. AEHR: <0.001<br>Active vs. Sedentary: 0.02<br>Sedentary vs. AEHR: <0.001 |

Abbreviations: AEHR, atypically elevated heart rate; QI, quadrant I; bpm, beats per minute; QII, quadrant II; QIV, quadrant IV.
